# Comparative Evaluation of rs-fMRI Representations for Differentiating Schizophrenia and Bipolar Disorder with Psychosis

**DOI:** 10.64898/2026.06.16.26355608

**Authors:** Daniela Janeva, Martin Breyton, Silvana Markovska-Simoska, Romain Guilhaumou, Spase Petkoski, Armin Iraji, Vince Calhoun, Branislav Gerazov

## Abstract

Psychosis is a prominent symptom dimension in both schizophrenia and bipolar disorder, two highly heterogeneous psychiatric disorders with partially overlapping clinical manifestations. Resting-state functional magnetic resonance imaging (rs-fMRI) represents a promising tool for identifying objective biomarkers of functional brain alterations to aid differential diagnosis. In this work, we comparatively evaluate multiple rs-fMRI representations for differentiating schizophrenia and bipolar disorder using intrinsic connectivity network (ICN) temporal profiles and several functional network connectivity (FNC) approaches, including static, dynamic, and high-order analyses. Models were developed on a labeled cohort of 471 subjects with psychosis (183 BP, 288 SZ) and evaluated on a separate held-out cohort of 315 subjects. We evaluated convolutional neural networks trained on ICN temporal profiles, spectrograms, and scalograms, together with classical machine-learning models trained on multiple functional network connectivity representations. Across the evaluated approaches, ICN temporal profiles provided the most consistent discriminative performance, with a 1D convolutional neural network achieving the strongest overall results under the benchmark protocol (test AUC = 0.671). Among connectivity-based methods, static sFNC generally outperformed dynamic and high-order representations, suggesting that increased representational complexity did not necessarily translate into improved generalization. Although the obtained classification performance remained modest, the results highlight the challenges of robust psychosis differentiation using rs-fMRI while emphasizing the relative stability of low-order connectivity representations and temporal ICN features. These findings contribute to ongoing efforts toward reproducible and interpretable neuroimaging biomarkers for psychiatric disorders.

## 1. Introduction

Schizophrenia (SZ) and bipolar disorder (BP) with psychosis are intricately intertwined psychiatric conditions that pose significant challenges in mental health diagnosis. Schizophrenia is identified by positive symptoms, including hallucinations and delusions, as well as negative symptoms such as social withdrawal, anhedonia, and avolition, alongside cognitive symptoms such as impaired attention and working memory (Andreasen and Flaum, 1991). In contrast, bipolar disorder (type 1) is marked by episodes of mania, featuring increased energy, racing thoughts, and risky behavior, followed by depressive episodes with symptoms including low mood, fatigue, appetite and sleep changes, as well as difficulty concentrating (Vieta et al., 2018).

Schizophrenia and bipolar disorder entail chronic symptoms that can severely impact daily life and functioning (Green, 2006; Murray et al., 2004). Although the two disorders are distinct clinical entities, they share similarities in symptoms and genetic risk factors, and both can involve cognitive deficits. This overlap has led some researchers to suggest that the two disorders may be part of a broader spectrum of mental illnesses that share underlying genetic and environmental risk factors (Maier et al., 2006). Accurate diagnosis is crucial, as misdiagnosis or delayed diagnosis can lead to inappropriate or ineffective treatments and poor patient outcomes. For instance, antipsychotic medications typically used for schizophrenia may exacerbate symptoms of mania in bipolar disorder, whereas mood stabilizers and antidepressants typically used for bipolar disorder may not effectively treat symptoms of schizophrenia (Narasimhan et al., 2007; Nayak et al., 2021).

Traditional diagnostic approaches, while valuable, may struggle to provide the discrimination necessary for effective clinical intervention. Despite extensive research, the underlying pathological mechanisms of schizophrenia and bipolar disorder remain partially unknown, with emerging evidence pointing to differences in brain connectivity as a potential key to understanding these conditions (Collin et al., 2016; Iraji et al., 2022b). Recent progress in neuroimaging has provided transformative insights into psychiatric conditions. Resting-state functional magnetic resonance imaging (rs-fMRI), in particular, has emerged as a promising tool to understand the underlying neural mechanisms of these disorders (Reavis et al., 2017). Rs-fMRI captures intrinsic neural activity by measuring low-frequency fluctuations in blood-oxygen-level-dependent (BOLD) signals, offering a window into the functional organization of the brain (Heeger and Ress, 2002).

Intrinsic connectivity networks (ICNs), obtained through independent component analysis (ICA) of resting-state fMRI data, portray coherent endogenous neural activity related to brain function. ICA decomposes rs-fMRI data into spatially independent components that represent distinct networks corresponding to different functional systems (Calhoun et al., 2001). ICNs have been extensively explored in psychiatric disorders (Calhoun et al., 2012; Iraji et al., 2022a). Functional network connectivity (FNC) is typically a measure of the linear relationship between ICN time courses, usually computed as Pearson’s correlation coefficient between each pair of ICN time courses. Static FNC (sFNC), which has been extensively studied, does not capture the complex spatiotemporal dynamics of the brain. Previous studies have consistently highlighted the significance of temporally changing interactions between brain regions in understanding disorders (Du et al., 2018; Hutchison et al., 2013). Dynamic functional network connectivity (dFNC), constructed through techniques such as the sliding-window approach, captures temporal fluctuations in brain network connectivity (Iraji et al., 2021). Additionally, high-order functional network connectivity (hFNC) explores interactions beyond conventional pairwise connectivity, providing a richer characterization of functional brain organization (Chen et al., 2016; Iraji et al., 2022b; Li et al., 2023).

In this work, we comparatively evaluate multiple rs-fMRI representations for differentiating schizophrenia and bipolar disorder with psychosis through two complementary analyses. In the first analysis, we investigate convolutional neural networks for classifying zero-padded ICN temporal profiles and their corresponding time–frequency representations, together with standard machine-learning algorithms applied to FNC computed from the full-length ICN time courses. In the second analysis, we standardize the ICN time courses to a fixed length to enable a fair comparison of static, dynamic, and high-order functional network connectivity (sFNC, dFNC, and hFNC) representations. Classification performance is evaluated using the area under the receiver operating characteristic curve (AUC) on an independent held-out test cohort. The remainder of this paper is organized as follows. Section 2 describes the dataset and methodologies. Section 3 presents and compares the experimental results, Section 4 discusses the findings, and Section 5 concludes the paper.

## 2. Materials and methods

The population sample used in this work consists of 786 patients in total. Of them, 183 BP and 288 SZ subjects are available for training, labeled correspondingly prior to this work, and 315 patients are reserved for testing.

The dataset includes time courses from 105 intrinsic connectivity networks estimated using the GIFT toolbox (Iraji et al., 2021), extracted from rs-fMRI using a multi-spatial-scale spatially constrained ICA template (Iraji et al., 2022c), and their corresponding functional network connectivity calculated as Pearson’s correlation between each pair of ICN time courses.

The provided features were extracted from the original datasets (Tamminga et al., 2013; Clementz et al., 2022) in several steps. Quality control was applied to identify high-quality data (Fu et al., 2024). The rs-fMRI data for each subject were pre-processed using a standard procedure, including rigid-body motion correction, slice-timing correction, and distortion correction following the NeuroMark pipeline (Du et al., 2020). Preprocessed subject data were registered into a common space, resampled to 3 mm^3^ isotropic voxels, and spatially smoothed using a Gaussian kernel with a 6 mm full width at half-maximum (FWHM). A multi-spatial-scale template of 105 ICs obtained from 100k+ subjects was used in a constrained ICA approach to obtain subject-specific ICN time courses. For calculating the FNCs, ICN time courses were cleaned using a common standard. FNC is generated by Pearson correlation between each pair of ICN time courses, resulting in one FNC matrix for each individual.

The methodological framework consists of two complementary analyses (Fig. 1). In the first analysis, we evaluated multiple rs-fMRI representations for differentiating schizophrenia and bipolar disorder with psychosis using full-length ICN time courses and their corresponding FNC. Specifically, we investigated deep learning models for ICN-based representations and classical machine-learning methods for full-length FNC features. In the second analysis, we standardized all ICN time courses to a fixed length to enable a fair comparison of static, dynamic, and high-order functional network connectivity (sFNC, dFNC, and hFNC) representations. For both analyses, the best-performing models were evaluated on an independent held-out test set using the area under the receiver operating characteristic curve (ROC-AUC). Soft probability scores on the test set were used to compute AUC.

**Figure 1.**
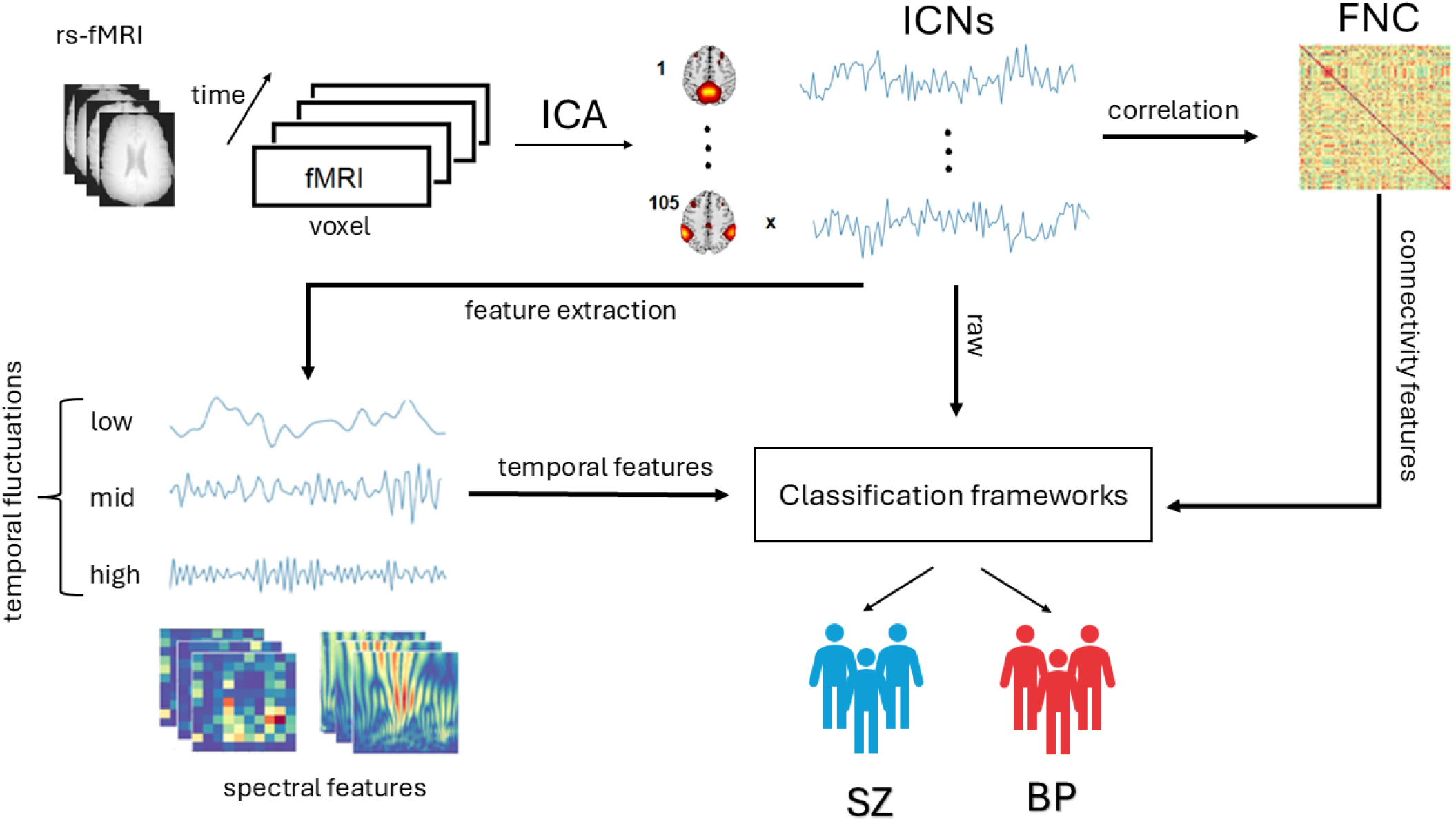
Overall workflow of the proposed study for differentiating schizophrenia and bipolar disorder using rs-fMRI-derived intrinsic connectivity networks (ICNs) and functional network connectivity (FNC) representations. The pipeline consists of preprocessing and feature extraction from ICN temporal profiles, followed by the generation of multiple representations, including filtered ICN signals, spectrograms, and scalograms. Deep learning architectures were employed for the classification of ICN temporal representations, while classical machine-learning models were used for connectivity-derived features. The resulting models were evaluated using soft probability predictions and area under the curve (AUC) scores on a held-out dataset.

### 2.1. Intrinsic Connectivity Network Temporal Profiles

Intrinsic connectivity, identified using fMRI, is a functionally interpretable relation of different brain regions (Laird et al., 2011), which is thought to reflect the underlying organization of the brain (Dovern et al., 2012). ICNs were identified using independent component analysis (ICA) (Calhoun et al., 2001; Allen et al., 2011). Significant alterations in ICNs in various neuropsychiatric disorders have been reported, emphasizing their importance for understanding the neural mechanisms underlying those conditions (Calhoun et al., 2021).

ICA models the observed multivariate fMRI signal **X** ∈ ℝ^*T* ×*V*^, where *T* is the number of time points and *V* is the number of voxels, as a linear mixture of *K* spatially independent components:

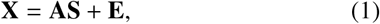

where **S** ∈ ℝ^*K*×*V*^ contains the independent spatial maps, **A** ∈ ℝ^*T* ×*K*^ represents the associated ICN time courses, and **E** is a residual error term (Calhoun et al., 2001; Allen et al., 2011).

The ICN time courses **A** capture the temporal evolution of spatially independent functional regions and provide a window into dynamic brain activity, which is particularly relevant in neuropsychiatric disorders such as schizophrenia and bipolar disorder (Calhoun et al., 2021). ICN time courses represent the temporal dynamics of ICA-derived spatial maps and serve as the primary temporal features in this study (*K* = 105).

#### Filtering

To homogenize differences in duration, we applied zero-padding to shorter time courses to align them with the maximum ICN length present in the dataset, which is 234 samples. We applied three bandpass Butterworth infinite impulse response (IIR) filters to obtain signals across three distinct frequency sub-bands: low, mid, and high. Assuming a sampling frequency of 0.5 Hz, the low-frequency band ranges from 0.075 to 0.125 Hz, the mid-frequency band from 0.125 to 0.175 Hz, and the high-frequency band from 0.175 to 0.25 Hz (Yuen et al., 2019). We divided the frequency range into low, mid, and high bands to explore ICN dynamics with balanced coverage of the spectral range.

#### Spectrogram and Scalogram

To derive the time–frequency representation (spectrogram) of the ICN time courses, we employed the short-time Fourier transform (STFT) using a sliding Tukey window of length 22 samples (Oppenheim and Schafer, 2010). The Tukey window provides a compromise between rectangular and Hann windows and was selected after empirical evaluation because it produced a clear time–frequency representation while balancing temporal and spectral resolution. Stacking spectrograms from ICN time courses resulted in a volumetric representation of dimensions 12 × 11 × 105, with frequency on the *x*-axis, time on the *y*-axis, and ICN index on the *z*-axis.

To obtain a representation with enhanced time–frequency resolution, we employed the continuous wavelet transform (CWT). The CWT offers an overcomplete decomposition using rescaled and translated replicas of a mother wavelet. The resulting scalograms preserve both temporal and spectral information and have been used as informative image representations for rs-fMRI analysis and deep learning (Al-Hiyali et al., 2021). We generated scalograms of the ICN components using the Morlet wavelet with 50 scales. Stacking the scalograms formed three-dimensional features of dimensions 49 × 234 × 105, where the *x*-axis denotes scales, the *y*-axis time, and the *z*-axis the IC index. Figure 2 illustrates exemplar raw and filtered ICN time courses, stacked spectrogram and scalogram tensors, and corresponding FNC matrices for representative SZ and BP subjects.

**Figure 2.**
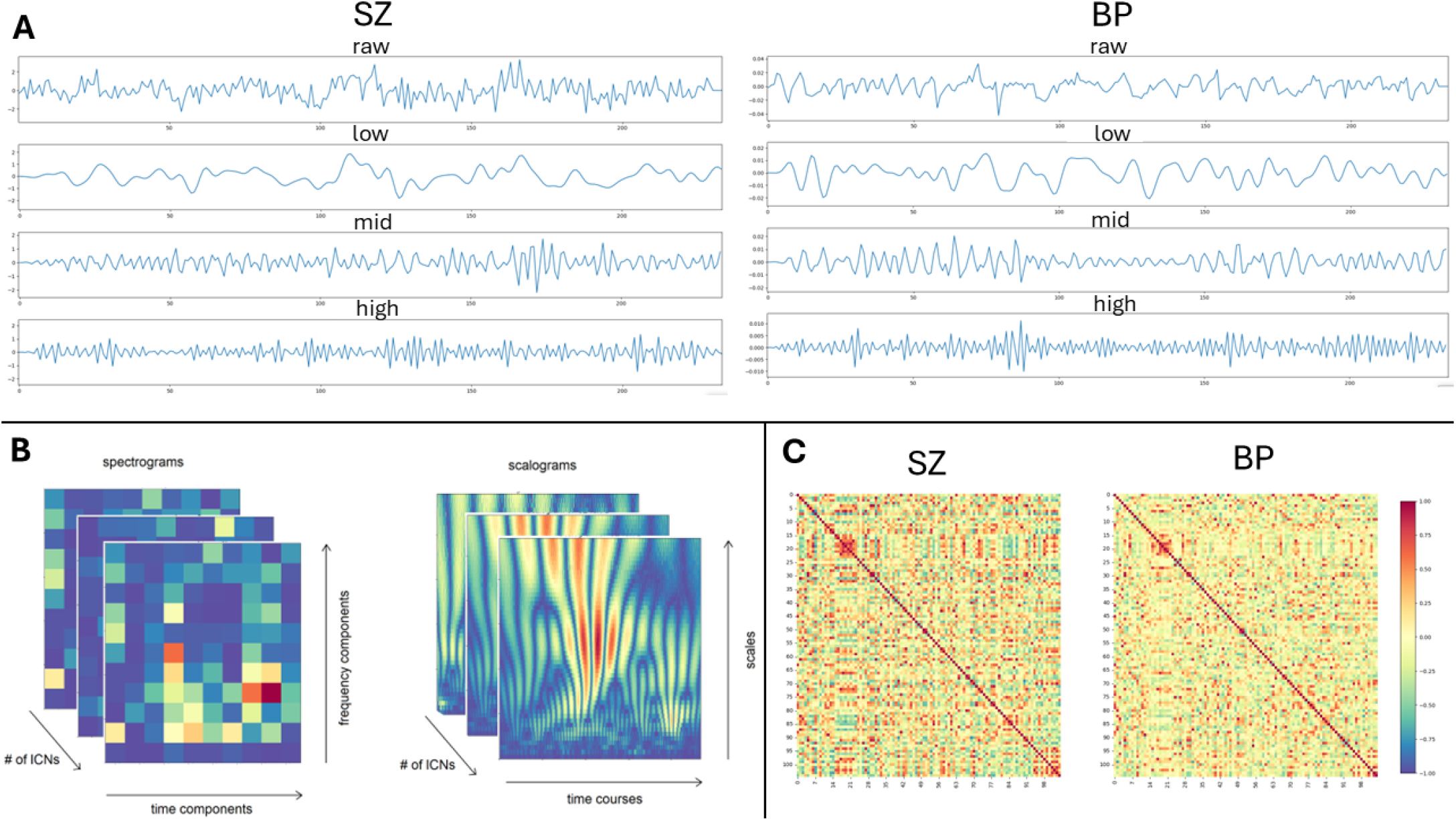
Sample IC component of an individual with SZ (left) and BP (right): raw signal, frequency sub-band signals, spectral representations, and functional network connectivity matrices. Panel A illustrates the raw and band-pass filtered ICN time courses for a representative subject with schizophrenia (left) and bipolar disorder (right), including the unfiltered raw signal and low-(0.075–0.125 Hz), mid-(0.125–0.175 Hz), and high-frequency (0.175–0.25 Hz) sub-bands. Panel B presents stacked spectrograms (12 × 11 × 105) and scalograms (49 × 234 × 105) across all 105 ICNs. Panel C displays the corresponding 105 × 105 FNC matrices.

#### Classification of Intrinsic Connectivity Network time courses

For classification of the raw ICN time courses and the sub-band signals obtained by filtering, we used a 1D convolutional neural network (CNN). The architecture is illustrated in Fig. 3A. Outputs from each convolutional layer are processed with a rectified linear unit (ReLU) activation function (Agarap, 2018). For classifying the 3D spectrogram and scalogram features, we used a 3D CNN model shown in Fig. 3B. For this step, we designed a simplified architecture by reducing the number of layers and trainable parameters due to computational constraints.

**Figure 3.**
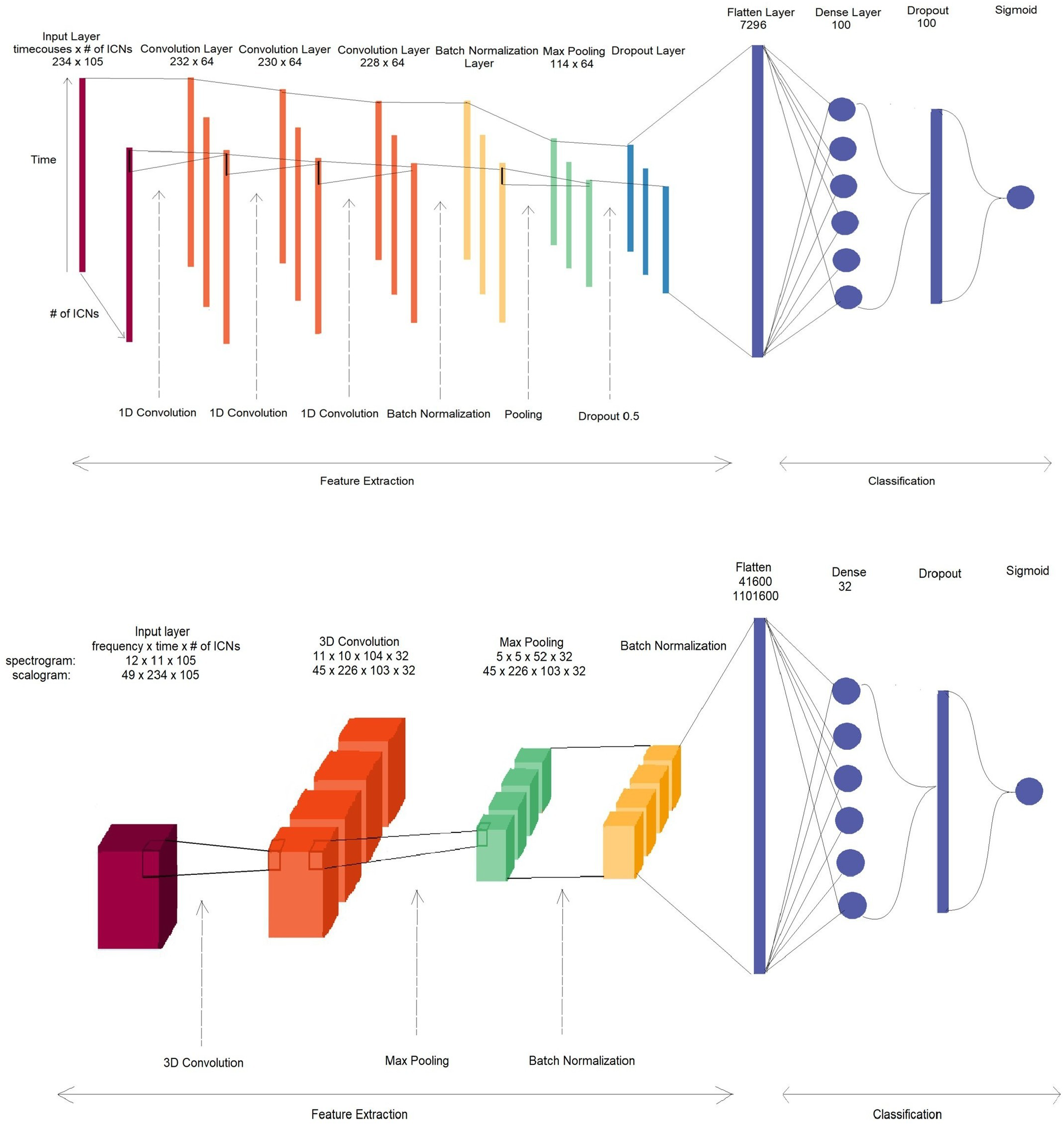
(A) 1D CNN used for raw and filtered ICN time courses. The input (234 ×105) passes through three 1D convolutional layers (64 filters, ReLU), followed by batch normalization, max pooling, and dropout (0.5), yielding a flattened vector fed into a dense layer (100 units) and a sigmoid output for binary classification. (B) 3D CNN used for spectrogram and scalogram features. The 3D input (12 × 11 × 105 or 49 × 234 × 105) is processed by a single 3D convolutional layer (32 filters), max pooling, and batch normalization, before a dense layer (32 units) and sigmoid output. This simplified architecture was adopted to accommodate the high dimensionality of the 3D inputs within available computational resources.

We trained both models using the Adam optimization algorithm (Kingma and Ba, 2014) for up to 100 epochs, with batch size 32. To prevent overfitting, we included early stopping with a patience of 20 epochs, monitoring binary cross-entropy on a validation set held out from the training data (Prechelt, 2002). The labeled dataset, consisting of 183 BP and 288 SZ patients, was randomly divided into 80% for training and 20% for validation, with no overlap between the subsets. The best-performing model, selected based on validation performance, was evaluated on an independent test set of 315 patients, and classification performance was quantified using the test-set AUC.

### 2.2. Functional Network Connectivity

Functional network connectivity represents patterns of synchronous activity between different brain regions that are postulated to underlie specific cognitive functions (Jafri et al., 2008). While traditional FNC calculations focus on distinct brain regions, in this study the concept is applied to ICN time courses, which represent temporal profiles of subnetworks. In that regard, our approach is closer to studies of FNC between resting-state networks (Rashid et al., 2016). In the dataset, FNCs are generated by calculating Pearson correlation coefficients between each pair of ICN time courses, which results in a symmetrical matrix of dimensions 105 × 105. The lower triangular matrix of the FNC is flattened into a vector of dimensions 1 × 5460.

#### Classification of Functional Network Connectivity

To classify individuals based on FNC features, seven classical supervised machine-learning algorithms were evaluated: support vector machine (SVM) with a radial basis function kernel, random forest (500 estimators), logistic regression, *k*-nearest neighbors (KNN; *k* = 5), Gaussian naïve Bayes, decision tree, and linear discriminant analysis (LDA). Each classifier was embedded within a standardized pipeline in which FNC features were *z*-score normalized using a StandardScaler fitted exclusively on the training set, with the same transformation applied to the held-out test set to prevent data leakage. Models were trained on the full training partition and evaluated on an independent test set. Model discrimination ability was assessed using ROC-AUC, computed from predicted class probabilities or decision-function scores. ROC-AUC served as the primary criterion for model selection given its robustness to class imbalance in clinical neuroimaging samples. Accuracy and F1 score were additionally computed for each model and visualized alongside ROC-AUC. The best-performing model was identified by highest ROC-AUC, and all random processes were seeded at 42 to ensure reproducibility.

### 2.3. Comparison of Static, Dynamic, and High-Order FNC

For static, dynamic, and higher-order FNC analysis, we used only the provided ICN time courses. We standardized the duration to a consistent length of 100 samples to match the shortest length in the dataset and computed each FNC from the standardized ICs. Standardization was necessary because dynamic and high-order connectivity require equal-length temporal inputs. Figure 4 summarizes the construction of these representations.

**Figure 4.**
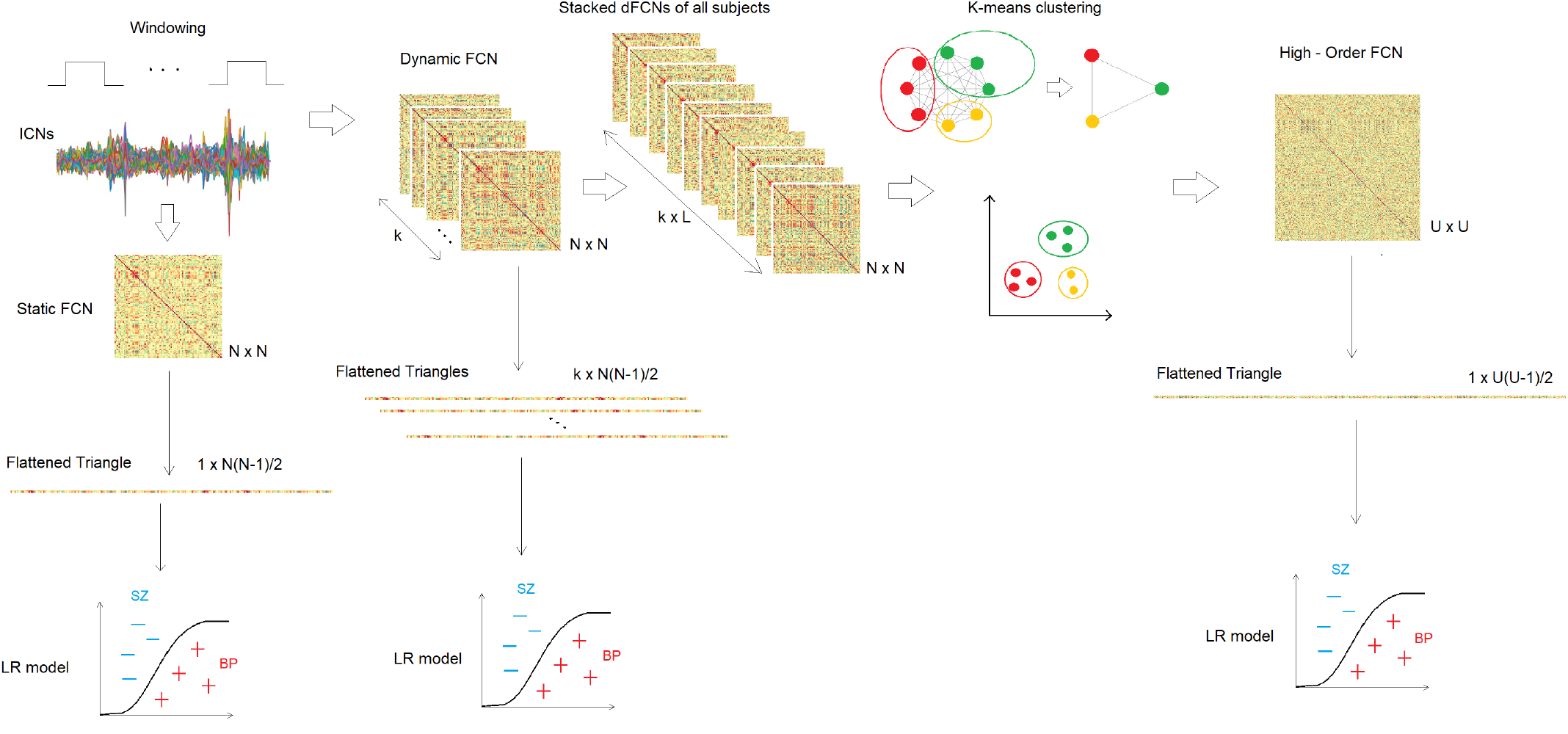
Methodology for length-matched connectivity features. Static FNC is computed as pairwise Pearson correlation of all ICN time courses; because of symmetry, the lower triangle is flattened and classified. Dynamic FNCs are calculated using sliding windows on the ICs; each window’s lower triangle is flattened and stacked. For higher-order FNCs, dFNC edge time series are stacked across subjects for unsupervised *k*-means clustering; cluster-mean series are correlated to obtain an hFNC of dimensions *U* × *U*, whose lower triangle is flattened for classification.

#### Static Functional Network Connectivity Analysis

FNC analysis is a widely used approach to study patterns of functional interactions between brain regions under the assumption that these patterns remain relatively constant over the recording (Liegeois et al., 2017; Allen et al., 2011). The static FNC provided in the dataset is computed for varying ICN time-course lengths. For each subject *l*, we computed the Pearson correlation between all pairs of ICN time courses standardized to a length of 100 samples, to enable a fair comparison across methods:

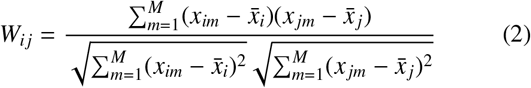

where *W*_*ij*_ = corr(*x*_*i*_, *x* _*j*_) is the Pearson correlation coefficient between ICN time courses *x*_*i*_ and *x* _*j*_ from **X** ∈ ℝ^*M*×*N*^, with *M* = 100 the length of the time courses and *N* = 105 the number of ICs for subject *l*. Here *x*_*im*_ and *x*_*jm*_ are the values of the time series for regions *i* and *j* at time point *m*, and 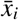 and 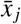 are the respective means. The resulting matrix **W** ∈ ℝ^*N*×*N*^ represents the sFNC for each subject. Due to symmetry, the lower triangle is flattened into a vector of length 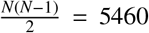 for classification.

#### Dynamic Functional Network Connectivity

Dynamic functional network connectivity aims to capture time-varying patterns of FNC. Unlike sFNC, which assumes constant connectivity over the recording, dFNC recognizes that network interactions change with time. In dFNC analysis, the time series are divided into overlapping segments using a sliding-window approach (Shakil et al., 2016; Allen et al., 2014).

The number of subseries *K* can be calculated as 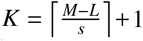, where *M* is the total duration, *L* is the window length, and *s* is the step size. Window length is a critical parameter: shorter windows capture rapid fluctuations but are more susceptible to noise, whereas longer windows capture more stable patterns but may overlook brief changes (Hindriks et al., 2016; Iraji et al., 2021). A smaller step size provides finer-grained updates on connectivity dynamics.

For the *l*-th subject, the *k*-th segment of all *N* ICN time courses is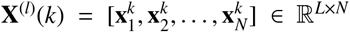. For each temporal FNC matrix with 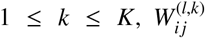 is computed as Pearson’s correlation as in Eq. (2). This yields *K* correlation matrices **W**^(*l*)^ ∈ ℝ^*N*×*N*^ and explores the temporal dynamics of intrinsic connectivity (Iraji et al., 2020). For classification, the lower triangle of each window matrix is extracted and stacked. Each correlation time series is 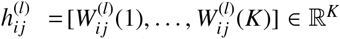. Stacking these series yields a matrix of dimensions *K* × 5460. In our experiments, we used combinations of *L* = [10, 20, 30, 40, 50] and *s* = [5, 10, 15, 20, 25], respectively.

#### High-Order Functional Network Connectivity

High-order FNC is obtained using the correlations-of-correlations principle (Pan et al., 2022). Computing relationships between dFNC time series captures high-order functional interactions across intrinsic networks (Chen et al., 2016). Each dFNC time series 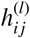 represents the temporal evolution of the Pearson correlation between the *i*-th and *j*-th ICN time courses for subject *l*. The high-order connectivity between pairs of dFNC time series is

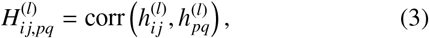

which quantifies how connectivity between ICNs *i* and *j* co-varies with connectivity between ICNs *p* and *q* over time. The number of high-order features scales as *O*(*N*^4^). For *N* = 105 ICNs, the number of high-order connections exceeds 10^6^. To reduce dimensionality, dFNC time series (lower-triangular entries for all subjects) were concatenated and *k*-means clustering was used to partition { *h*_*ij*_ *}* into *U* clusters. The mean dFNC time series within each cluster was then used to construct a compact hFNC representation.

The clustering algorithm assigns *h*_*ij*_ with similar temporal variation to the same cluster Ω_*u*_, where 1≤ *u* ≤*U*. The mean correlation series for patient *l* from cluster *u* is

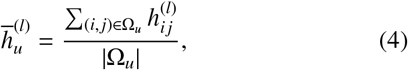

where |Ω_*u*_| is the number of elements in the cluster. High-order connectivity is then 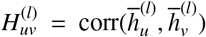. This reduces complexity from *N*^4^ to *U*^2^. The lower triangle of 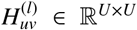 is flattened to length *U*(*U −* 1)*/*2. For each pair of *L* = [10, 20, 30, 40, 50] and *s* = [5, 10, 15, 20, 25], hFNCs were calculated for *U* = [10, 50, 100, 200, 300].

#### Classification of sFNC, dFNC, and hFNC

We used a logistic regression model for differentiating schizophrenia and bipolar disorder using each of these features (Pedregosa et al., 2011), whose extraction steps are shown in Fig. 4. We trained the models and optimized hyperparameters by 5-fold cross-validation over a parameter grid with grid search. For each experiment, the best models were used to predict soft probability scores on the held-out test dataset, and evaluation was performed using AUC as described previously.

## 3. Results

### 3.1. Classification Performance of ICN and Full-Length FNC Representations

The first analysis evaluated the discriminative performance of ICN-based representations and full-length FNC features for differentiating schizophrenia and bipolar disorder with psychosis. The AUC scores from our CNN models for raw and filtered ICN time courses are shown in Fig. 5. The best score was obtained by the 1D CNN trained on raw IC time courses (test AUC = 0.671). Among the band-limited representations, the low-frequency sub-band (0.075–0.125 Hz) yielded AUC = 0.546, the mid-frequency sub-band (0.125–0.175 Hz) AUC = 0.54, and the high-frequency sub-band (0.175–0.25 Hz) AUC = 0.53. Classification of scalograms produced a better AUC than spectrograms, but both were weaker than raw ICN inputs overall. The 3D CNN achieved an accuracy of 0.63 and AUC of 0.53 for scalogram classification, and an accuracy of 0.60 and AUC of 0.49 for spectrogram classification.

**Figure 5.**
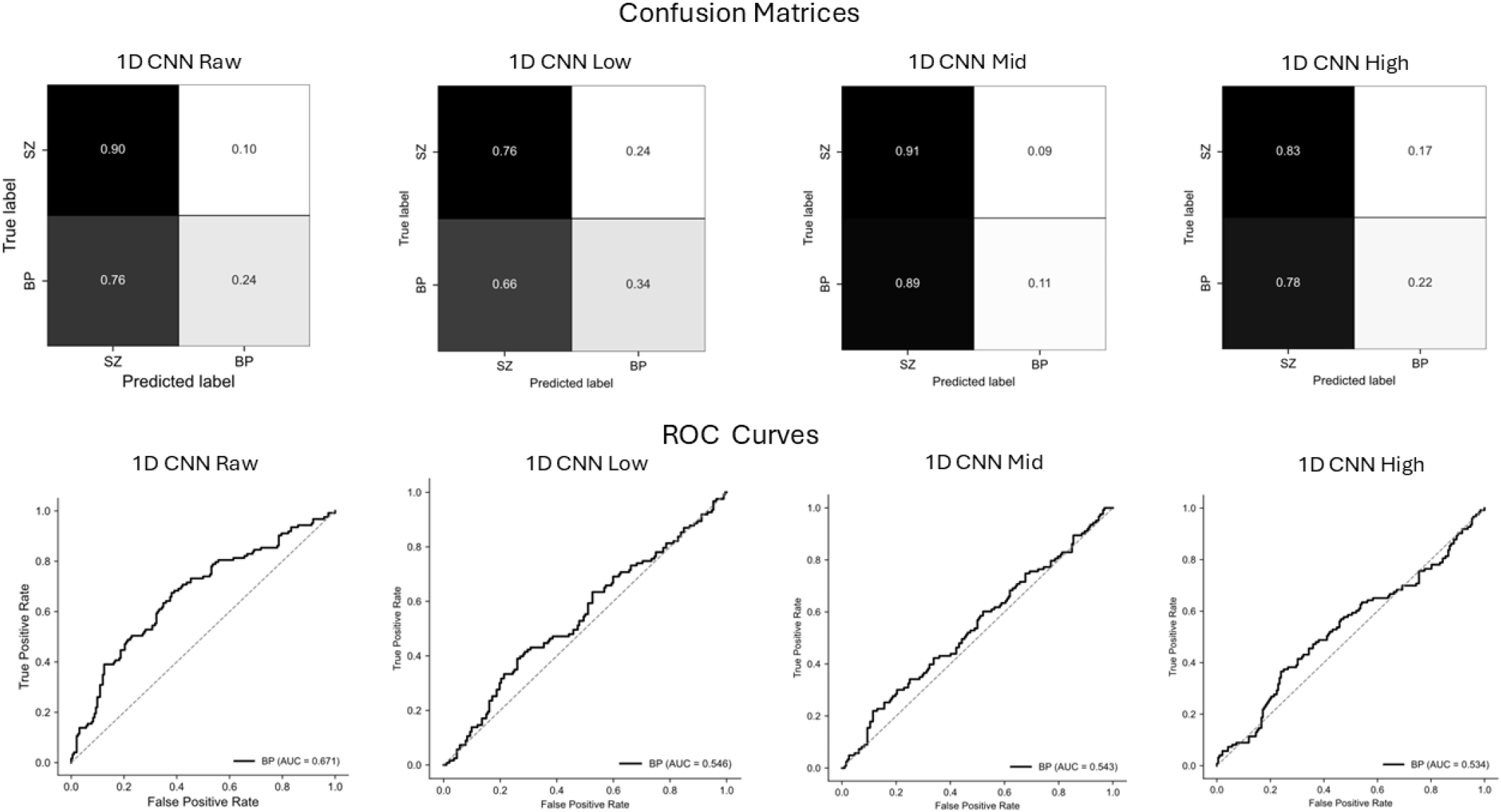
Confusion matrices (top row) and receiver operating characteristic (ROC) curves (bottom row) for the 1D CNN classifier evaluated using raw signals and three frequency-band representations (low, mid, and high). The confusion matrices show the normalized classification performance between schizophrenia (SZ) and bipolar (BP) classes, with values representing the proportion of predictions per true class. ROC curves illustrate the trade-off between true positive rate and false positive rate for BP classification, with corresponding AUC values indicated in each panel. Among the evaluated inputs, the raw-signal model achieved the highest discriminative performance (AUC = 0.671), whereas the band-limited representations showed comparatively lower classification performance.

Among the seven classical machine-learning classifiers evaluated on FNC features generated by correlating full-length signals, logistic regression achieved the highest ROC-AUC of 0.640, followed by SVM (0.632) and LDA (0.620), as shown in Fig. 6. The decision tree classifier performed at near-chance level (AUC = 0.502). Random forest achieved the highest raw accuracy (~0.68) but exhibited a markedly low F1 score of approximately 0.20, indicating substantial class imbalance in its predictions. Most classifiers demonstrated moderate, consistent performance across accuracy, F1, and ROC-AUC, with F1 scores generally lagging behind the other two measures.

**Figure 6.**
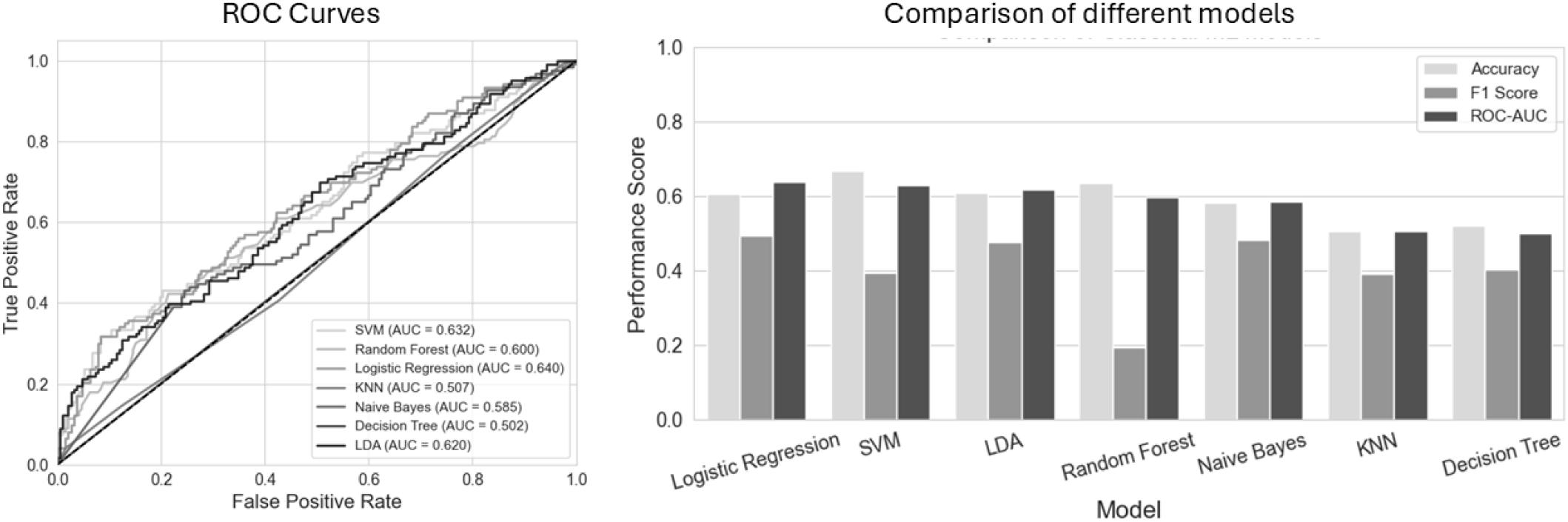
Performance comparison of seven classification models. (Left) ROC curves for all models, with logistic regression achieving the highest AUC (0.640), followed by SVM (0.632) and LDA (0.620). Decision tree performed poorest (AUC = 0.502), near the level of random chance. (Right) Bar chart comparing accuracy, F1 score, and ROC-AUC across models. Random forest achieved the highest accuracy (~0.68) but exhibited a markedly low F1 score (~0.20), suggesting poor precision–recall balance. Most models showed moderate and consistent performance across all three metrics, with F1 scores generally lagging behind accuracy and ROC-AUC. Abbreviations: SVM, support vector machine; KNN, *k*-nearest neighbors; LDA, linear discriminant analysis; ROC-AUC, receiver operating characteristic–area under the curve.

### 3.2. Comparison of Static, Dynamic, and High-Order Functional Connectivity Representations

Table 1 presents the AUC scores obtained by classifying low-order FNC representations derived from shortened ICN time courses. Static FNC achieved the highest AUC of 0.570, marginally outperforming all dFNC configurations. Among the dFNC settings, the shortest window (*L* = 10, *s* = 5) yielded the best result (AUC = 0.569), with performance declining as window length increased to a minimum of 0.548 at *L* = 30, *s* = 15, before slightly recovering at larger windows. It is important to note that shortening the ICN time courses prior to computing sFNC yielded worse AUC scores compared with previously reported full-length results (Janeva et al., 2023). Therefore, longer rs-fMRI sessions appear important for accurate differentiation. Overall, FNC classification demonstrated poorer performance than classification of ICN time courses under the present protocol.

**Table 1:** Classification performance of static and dynamic functional network connectivity representations computed from shortened ICN time courses.

| Network, Parameters | AUC |
| --- | --- |
| sFNC | 0.570 |
| dFNC $L = 10, s = 5$ | 0.569 |
| dFNC $L = 20, s = 10$ | 0.556 |
| dFNC $L = 30, s = 15$ | 0.548 |
| dFNC $L = 40, s = 20$ | 0.564 |
| dFNC $L = 50, s = 25$ | 0.564 |

Static (sFNC) and dynamic (dFNC) functional network connectivity matrices were computed from ICN time courses standardized to a fixed length. Performance is reported as ROC-AUC on the held-out test set.

Table 2 reports the AUC scores for hFNC across varying numbers of clusters (*U*) and window/step configurations. Performance across all hFNC configurations was consistently lower than both sFNC and dFNC, with scores ranging from 0.436 to 0.565. No consistent trend was observed with respect to either the number of clusters or the window parameters, suggesting that the high-order representations did not capture diagnostically discriminative information in a stable or predictable manner under the current experimental setup. The choice of window length, step size, and number of clusters was not coherently related to AUC; further adjustments or nested optimization of these parameters could potentially improve performance.

**Table 2:** Classification performance of high-order functional network connectivity across different clustering and window parameters.

| Parameters | $U = 10$ | $U = 50$ | $U = 100$ | $U = 200$ | $U = 300$ |
| --- | --- | --- | --- | --- | --- |
| $L = 10, s = 5$ | 0.521 | 0.478 | 0.479 | 0.520 | 0.514 |
| $L = 20, s = 10$ | 0.565 | 0.474 | 0.485 | 0.531 | 0.459 |
| $L = 30, s = 15$ | 0.495 | 0.517 | 0.455 | 0.513 | 0.508 |
| $L = 40, s = 20$ | 0.436 | 0.480 | 0.444 | 0.494 | 0.524 |
| $L = 50, s = 25$ | 0.522 | 0.475 | 0.524 | 0.558 | 0.535 |

High-order functional network connectivity (hFNC) was computed from dynamic FNC derived from ICN time courses standardized to 100 samples. Rows indicate the sliding-window length ( $L$ ) and step size ( $s$ ), whereas columns correspond to the number of $k$ -means clusters ( $U$ ). Values represent ROC-AUC on the held-out test set.

## 4. Discussion

The results of this study demonstrate that the proposed ICN-based classification pipeline, particularly when using raw ICN time courses processed with a 1D convolutional neural network, achieved the highest classification performance among the evaluated approaches (AUC = 0.671). Because different classification models were employed for ICN-based and connectivity-based representations, these findings should not be interpreted as evidence that ICN temporal profiles are inherently more discriminative than functional connectivity features. Rather, they indicate that the proposed ICN-based framework provided the best performance under the evaluated experimental setting. The superior performance of raw time courses over filtered sub-band signals suggests that diagnostically relevant information is distributed across the full frequency spectrum rather than being confined to any single sub-band, and that decomposing the signal into narrow frequency bands may discard complementary cross-band dynamics.

The underperformance of 3D CNN architectures on spectrogram and scalogram features is likely attributable to the increased dimensionality of the input combined with the limited size of the available training cohort. The high dimensionality of 3D representations introduces a substantial risk of over-fitting that the simplified architecture, adopted due to computational constraints, may not have been sufficient to mitigate.

Future work employing larger datasets or more aggressive regularization strategies may reveal greater utility in these time– frequency representations.

Within the connectivity-based analysis, length-matched sFNC matched or exceeded the sliding-window dFNC and cluster-compressed hFNC representations evaluated here. This does not imply that dynamic or high-order connectivity lacks clinical value; rather, under the present length-standardized protocol, these specific constructions did not improve SZ–BP discrimination relative to static FNC. Recent work continues to show that transient network dynamics can encode clinically meaningful information in schizophrenia, suggesting that alternative dynamic or high-order representations may succeed where the conventional sliding-window and correlations-of-correlations setup used here did not (Iraji et al., 2024; Li et al., 2023). A further contributing factor is that truncating ICN time courses to 100 samples reduces the temporal support available to dFNC and hFNC. Consistent with this, full-length sFNC out-performed the shortened sFNC, underscoring sensitivity to acquisition duration.

The lack of a consistent relationship between hFNC parameters and classification performance suggests that optimization of high-order connectivity remains challenging under the current experimental conditions. Systematic hyperparameter optimization using nested cross-validation, or the adoption of data-driven clustering strategies, may help identify configurations that yield more stable and interpretable high-order features. Nevertheless, the overall modest performance across all methods reflects the fundamental difficulty of the task: schizophrenia and bipolar disorder share substantial genetic, neurobiological, and symptomatic overlap, and the rs-fMRI signal, while informative, may not carry sufficient disorder-specific variance to enable robust differentiation in cohorts of this size.

The training set exhibited a moderate class imbalance (288 SZ vs. 183 BP subjects), which may have contributed to the lower F1 scores observed for some classifiers, particularly Random Forest. Future work may benefit from exploring class-balancing strategies such as oversampling or cost-sensitive learning.

## 5. Conclusions

We compared ICN temporal representations and static, dynamic, and high-order connectivity features for differentiating schizophrenia and bipolar disorder with psychosis on an independent test cohort. Raw ICN time courses classified with a 1D CNN provided the best discrimination in this study, while full-length static FNC was the strongest connectivity baseline. Increasing representational complexity via filtering, time–frequency transforms, or high-order connectivity did not improve generalization under the evaluated protocol. These findings support prioritizing robust low-order temporal and static connectivity features—and adequate scan duration—in future biomarker work, while underscoring that current rs-fMRI pipelines alone are not yet sufficient for reliable clinical differentiation.

## Data Availability

All data produced in the present study are available upon reasonable request to the authors

